# Estimating economies of scale and learning-by-doing effects in a health systems strengthening intervention

**DOI:** 10.1101/2025.06.17.25329775

**Authors:** Lori A Bollinger, Kristin Bietsch

**Affiliations:** Avenir Health, Glastonbury, CT, USA

## Abstract

It can be challenging to evaluate efficiencies for interventions strengthening health systems, for a variety of reasons. As part of a program review of The Challenge Initiative (TCI) that took place in 2020, we explored whether economies of scale and learning-by-doing effects existed for the health systems strengthening portion of the project, utilizing audited expenditure data and validated reported outcomes. TCI is a project that partnered with local governments in low- and middle-income countries to deliver interventions for family planning; it is relatively unique in terms of both its activities and in how its expenditures and outcomes were tracked. Using TCI data, we estimated a cost function, evaluating economies of scale using as output measures both the number of geographies that joined the project over time and the population of women of reproductive age in those geographies over time, allowing for complex effects by including levels, squared and cubic formulations of the output variables. We also evaluated whether learning-by-doing effects obtained by examining coefficients of time-related variables. Results suggested that while initially there were diseconomies of scale when output was measured using number of geographies, the TCI program began to experience economies of scale as the number expanded, with significant economies of scale experienced beginning at the mean number of geographies. When output was measured using population of women of reproductive age, economies of scale existed throughout. We did not find econometric evidence of learning-by-doing effects.

## Introduction

It can be challenging to evaluate efficiencies for interventions strengthening health systems for a variety of reasons, although a few examples exist in the literature ([1], [2], [3]): the difficulty of separating out specific health systems strengthening expenditures from overall expenditures; challenges with identifying appropriate outcomes and then measuring those outcomes over time; and reluctance of implementers to share data with researchers who could perform efficiency analyses. The lack of this research is particularly prevalent in low-and middle-income countries; a recent systematic review identified only 36 studies that performed any economic evaluations at all of health systems strengthening efforts in low- and middle-income countries [4]; note that none included quantitative evaluations of efficiencies. Instead, the studies reported very specific results, leading the authors to recommend using techniques going forward to perform analyses that are more generalizable, such as estimating cost functions. This recommendation is echoed by other authors in a recent methodological paper, which defined a suggested typology of health systems strengthening interventions and then discussed illustrative examples of evaluations that would be useful, including efficiency analyses [5].

The Challenge Initiative (TCI) is a project funded by the Gates Foundation that partners with local governments for geographies (including cities, counties, districts, and states) in low- and middle-income countries to deliver high impact interventions (HII) for family planning. Local governments could choose from amongst a range of HIIs, including improving the management of FP programming, supporting advocacy to increase funding for FP, improving service delivery, and generating demand. The local governments are then supported by TCI through several sets of activities including accessing toolkits that guide the HII implementation; coaching to strengthen managerial and technical capacity of local government staff, service providers, and community health workers; and participating in a community of practice to brainstorm ideas and share lessons learned (see <u>TCI University</u> for further details). Funds pledged by local government are then matched by program funds from Gates Foundation and others to implement the selected HIIs; note that TCI did not fund the provision of family planning services.

As part of a 2020 program review, funded by Gates Foundation, we explored whether economies of scale and learning-by-doing effects existed for the health systems strengthening portion of the project, utilizing audited expenditure data and validated reported outcomes. TCI was relatively unique in terms of both its activities in the health systems strengthening sphere and in how its results were tracked, resulting in high-quality, reliable data both on expenditures related specifically to health systems strengthening, and on the output measures used to evaluate the project, addressing two of the challenges to performing efficiency analyses noted above. Further, since the research was being conducted as part of a program review, researchers were given full access to both the data and to TCI staff who clarified issues and answered questions that arose.

Below, we describe the data we utilized and the methodological approach we followed to evaluate whether economies of scale and/or learning-by-doing effects were found in the TCI program review. We then present the results of the evaluation, finishing with a discussion section that describes the implications of the results for TCI as well as implications for the field more broadly.

## Methods

Economies of scale (EOS) occur as fixed costs are spread out over larger and larger quantities of output, illustrated in Figure 1 by a movement along the average cost curve AC1 as indicated by the black arrow; note that the production technology underlying the average cost curve itself does not change. The extent to which significant efficiencies can be obtained through increasing the scale of production depends on the magnitude of the fixed costs. On the other hand, learning-by-doing effects are an actual change in technology, shifting the entire average cost curve downwards from AC1 to AC2, as indicated by the green arrow in Figure 1. Learning-by-doing effects are a change in technology because at every point along the average cost curve the cost is lower due to higher productivity levels. For TCI, economies of scale could have resulted from spreading fixed management costs as well as fixed design costs over the various implementation levels: headquarters vs. hub vs. TCI geography. However, declines in cost per unit could also have resulted from learning-by-doing effects, as staff learn to perform their jobs more efficiently over time. Because the two effects have the same result - lowering the cost per output – separating them required estimating a cost function.

**Figure 1:**
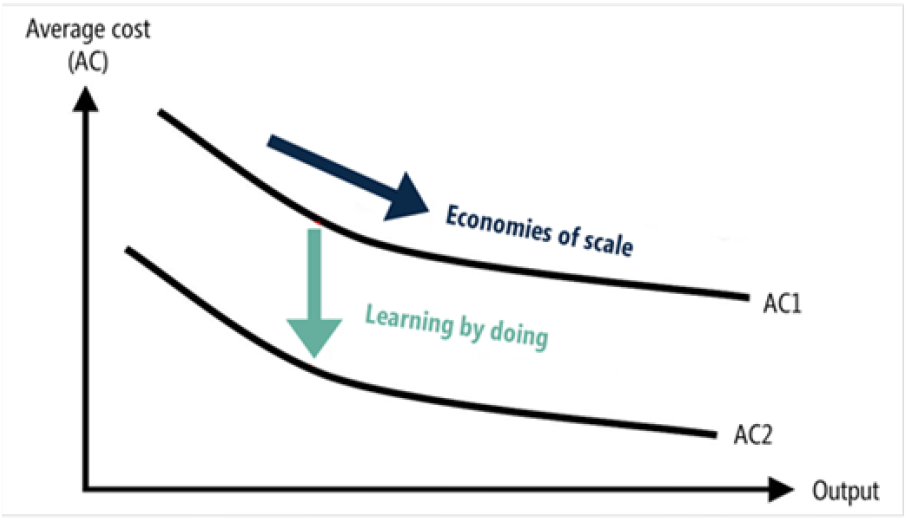
Economies of scale and Learning-by-doing effects.

We adapt previous work [6], [7] performing econometric analyses to assess potential efficiency gains by estimating a cost function where total costs are a function of input costs (personnel and all other costs) and two measures of quantity output (number of geographies per hub and relevant number of women of reproductive age (WRA)) in addition to other variables that might affect output, including regional dummy variables and a time trend to account for technological changes, including learning-by-doing effects. TCI provided us with quarterly expenditures for four years disaggregated into personnel and all other costs associated with providing the health systems strengthening components of TCI for the global^1^- and hub-level (East Africa, French West Africa (FWA), India, and Nigeria), including developing the materials for TCI-U, performing the coaching, and facilitating the community of practice. In addition, TCI provided us with the number of geographies that participated in the program for each of the corresponding quarters for each hub as well as the number of women of reproductive age (WRA) associated with each geography. ^2^

We assumed that the cost function is well-behaved mathematically (i.e., linearly homogeneous in input prices), and that TCI behaved in a cost-minimizing way, such that the cost function is:

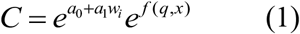

where: C = average total cost; a_0_ = constant; a_i_ = coefficients associated with i input prices; w_i_ = vector of inputs; q = number of geographies/WRA; and x = vector of independent variables that might shift the cost function. Taking the logarithm of both sides and including linear, squared, and cubed output (i.e., number of geographies/WRA) variables in the specification results in the following equation to be estimated:

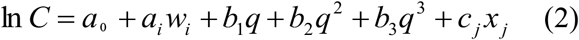

where b_i_ are the coefficients associated with the three output variables. To derive a measure of EOS, we calculate the marginal cost (MC, or *∂C /∂q*) by differentiating C in equation (2) with respect to q:

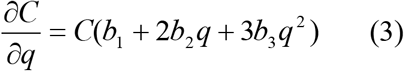

The EOS measure, which is an indication of efficiency, can then be calculated as:

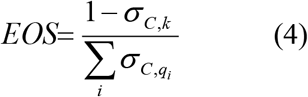

where *σ* _*a,b*_ is the elasticity of a with respect to b, and k is capital stock. Note that 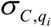 is equal to the product of the marginal cost of output (equation (3)) and the ratio of output to total cost. Since capital stock is a minimal part of the program, any variations are also minimal, and we can assume that the elasticity of substitution with respect to capital,*σ*_*C,k*_, equals zero [7]. Thus, as we have only one output q, the equation to calculate economies of scale becomes:

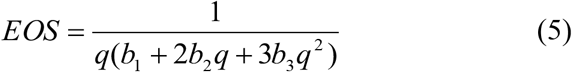

When EOS equals one, there are constant returns to scale, i.e., when inputs are doubled the resulting level of outputs is also doubled, if input prices remain constant. When EOS is greater than one, the level of output is less efficient, i.e., production could be more efficient if the level of output increases. When EOS is less than one, the level of output is more efficient.

Note that the coefficient on the time trend will be a proxy for learning-by-doing effects; we expect the coefficient to be negative, i.e., learning-by-doing should mean that costs decrease over time, although the coefficient could be conflated with other technological changes that have a positive effect if they are present.

We used Stata version 16 to perform the econometric estimation using a generalized linear model (GLM), correcting for heteroskedasticity by calculating robust standard errors using the Huber-White Sandwich estimator.^3^ We explored several specifications, including adding dummy variables for each year of the TCI program instead of a single time variable, capturing whether there were year-specific effects related to costs (e.g., significant start-up costs), as well as an interaction term for time and output (linear only), capturing whether there might be an accelerated effect for learning-by-doing as output increases.

## Results

Final regression results for the two specifications, each using a different output variable, are shown in Table 1. All independent variables were statistically significant at the 10% level or greater except for some of the regional dummy variables (the excluded regional variable is global). The coefficients for the input categories are all positive, which is expected *a priori*, and highly statistically significant. The coefficients for all three output terms (linear, squared, and cubed) are statistically significant for both output measures, as are both the Time variable alone and the interaction term of Time x output variable.

**Table 1:** Regression results from econometric estimation of cost function.

| Dependent variable:<br>Ln(Total costs) | Specification #1: Geographies |  | Specification #2: WRA |  |
| --- | --- | --- | --- | --- |
|  | Coefficient | P>z | Coefficient | P>z |
| Personnel | 1.26E-07*** | 0.002 | 1.55e-07*** | 0.000 |
| All other costs | 1.08E-07*** | 0.000 | 9.85e-08*** | 0.000 |
| Time | 0.00675** | 0.032 | .005721** | 0.014 |
| East Africa | -0.0126 | 0.222 | -.0196 | 0.171 |
| FWA | -0.0306* | 0.090 | -.0154 | 0.309 |
| India | -0.051** | 0.042 | -.0379 | 0.105 |
| Nigeria | -0.0533** | 0.075 | -.0373* | 0.064 |
| Geographies | 0.00412** | 0.044 |  |  |
| Geographies squared | -0.0000961* | 0.058 |  |  |
| Geographies cubed | 8.23E-07** | 0.016 |  |  |
| WRA |  |  | 2.03e-08*** | 0.007 |
| WRA squared |  |  | -1.79e-15** | 0.015 |
| WRA cubed |  |  | 5.81e-23*** | 0.010 |
| Time x Geographies |  |  |  |  |
| Time x WRA |  |  | -9.14e-10*** | 0.005 |
| Constant | 2.471*** | 0.000 | 2.459*** | 0.000 |
| Specification #1 (Geographies): n=75; AIC=7.47; BIC=-277.00; Specification #2 (WRA): n=76; AIC=7.39; BIC=-290.00<br>*** significant at 1% level; ** significant at 5% level; * significant at 10% level |  |  |  |  |

These results can then be used to calculate both learning-by-doing effects and economies of scale. We evaluate both at the mean, first quartile and third quartile values of the relevant independent variables (see Table 2); note that the calculations include consideration of the partial derivatives for the interaction Time x Geographies/WRA term for both learning-by-doing effects and economies of scale.

**Table 2:** Relevant statistics and calculation of learning-by-doing effects and economies of scale.

|  | Time | Output | Learning-by-doing effect | Economies of Scale |
| --- | --- | --- | --- | --- |
| Output=Geographies |  |  |  |  |
| Mean | 9 | 21 | 0.00237 | -66.41 |
| Quartile 1 | 5 | 2.5 | 0.00624 | 16.87 |
| Quartile 3 | 13 | 31 | 0.00040 | -15.14 |
| Output=WRA |  |  |  |  |
| Mean | 9 | 5,714,096 | 0.00050 | -67.43 |
| Quartile 1 | 5 | 564,179 | 0.00521 | 0.0001 |
| Quartile 3 | 13 | 6,841,228 | -0.00053 | -18.47 |

Recall that learning-by-doing effects are present when the overall effect is negative, i.e., costs decrease when learning-by-doing effects are present. Here, when Geographies is the output variable, the negative coefficient on the Time x Geographies term (see Table 1) is not large enough to outweigh the positive coefficient on the Time term alone when evaluating the overall learning-by-doing effects at the mean, first quartile or third quartile levels, resulting in positive effects for all three, i.e., an upward shift of the average cost curve. When WRA is the output variable, the overall effect is also positive at the mean and first quartile levels but does become slightly negative at the third quartile.

The calculations for economies of scale indicate that, when the number of TCI geographies is the output variable, initially (i.e., evaluated at the first quartile) there are potential economies of scale to exploit (EOS > 1) but then production does become efficient (EOS<1) at both the mean and third quartile, with some amelioration at the third quartile. When the output variable is number of WRA in the TCI geographies, economies of scale exist across the curve, although the effect again decreases somewhat by the third quartile.

## Discussion

As part of a program review of TCI, we estimated a cost function to evaluate whether expenditures associated with the health systems strengthening component of the program exhibited economies of scale and/or learning-by-doing effects. We were able to overcome some typical challenges faced when estimating efficiencies of health systems strengthening interventions: by design, TCI’s health systems strengthening component of the overall program was well-defined, with corresponding audited financial data; the definition and measurement of output variables was straightforward and the data validated; and we had unfettered access to records and TCI staff as the exercise was part of the program review.

There are several key findings from this research. First, for the most part, learning-by-doing effects do not appear to have been present in the TCI project. Except in one instance, while there is some ameliorating impact on the positive coefficient on the Time term alone when the interaction term is included, overall the calculations for the learning-by-doing effect are positive, i.e., implying an upward shift in the average cost curve, which is the opposite direction than was expected *a priori*. The one exception is when the learning-by-doing effects are evaluated at the third quartile using WRA as the output variable; there the overall effect is negative, implying that learning-by-doing occurs as the population served increases, i.e., the average cost curve shifts downward. One question that might affect this conclusion is whether other technological changes are present, conflating potential learning-by-doing effects; as part of the evaluation, we conducted extensive key informant interviews, during which we explored whether there were any other changes in technology that might affect results. TCI staff stated that the only country for which there might have been conflating factors would be Kenya, which was part of an earlier grant, the Urban Reproductive Health Initiative. However, TCI staff felt that since TCI was implementing a completely different set of interventions, i.e., was an entirely new model, the learning-by-doing effects estimated here could be relied upon to represent learning-by-doing effects due to the TCI project only. Other technological changes, such as changes in internet capabilities or access, are likely unchanged over the short evaluation time horizon of the TCI program.

Second, the TCI program does appear to have experienced economies of scale either across the entire average cost curve, in the case when output was measured by number of WRA, perhaps due to its higher variance, or as it moved towards the mean and higher output levels when the output variable is number of TCI geographies. The curvature is quite pronounced for both output variables, with the EOS parameter evaluated at the first quartile being relatively low (and exhibiting potential for further efficiencies as the scale of production increases when the output variable is TCI geographies), moving towards significant EOS at the mean, and then moderating to a lower level of EOS by the third quartile.

Third, we want to note that the regression results were extremely robust. When we first suggested trying to evaluate efficiencies as part of the program review by estimating a cost function, including separating out the two effects of learning-by-doing and EOS, we were very skeptical about whether it would even be possible. We were quite pleasantly surprised – and very happy – to discover that the regression results were very consistent and highly statistically significant across all specifications, including for the squared and even the cubic terms for the output variables.

One of the limitations of this analysis is whether one of the assumptions required to estimate a cost function, that the entity acts in a “cost-minimizing way,” holds here. TCI is an expensive project; the program review calculated that the average cost per additional user of modern contraception was $97 and the average cost per couple year of protection (CYP) was $44; note that this cost does not include commodities or service delivery costs, as TCI does not perform service delivery, thus the full cost per additional user/CYP would be even higher. Also, note that the coefficients for the hub dummy variables are all negative, suggesting that the hub expenditures were uniformly lower than global expenditures. Thus, results may not be entirely reliable if TCI was not acting in a cost-minimizing way.

Overall, however, given the ease and robustness of results which we experienced in the estimation process, we do recommend that others attempt to estimate EOS for health systems strengthening interventions. For this to occur, our second recommendation is that other projects build the necessary reporting into their project, including categorizing expenditures correctly and reporting outcomes accurately, so that this part of the program evaluation is built into their project design. If the expenditures and results of the health systems strengthening components of an intervention can be identified, and the data made available, then the estimation process will likely proceed in the same relatively seamless way we enjoyed here.

## Data Availability

All data produced in the present study are available upon reasonable request to the authors

## Acknowledgements

The research in this paper was funded by the Bill and Melinda Gates Foundation (INV-016714).

## Footnotes

1 The global level refers to the William H. Gates Institute Sr. for Population and Reproductive Health at Johns Hopkins Bloomberg School of Public Health, which leads TCI and supervises four subcontractors, one for each of four regional hubs East Africa, French West Africa (FWA), India, and Nigeria.

2 The number of geographies/WRA for the global level was defined as the sum of all geographies/WRA.

3 The actual STATA command used was: glm depvar indepvars, link(log) family(gamma) vce(robust).

